# A multi-ancestry genome-wide association study in type 1 diabetes

**DOI:** 10.1101/2023.09.18.23295708

**Authors:** Dominika A. Michalek, Courtney Tern, Wei Zhou, Catherine C. Robertson, Emily Farber, Paul Campolieto, Wei-Min Chen, Suna Onengut-Gumuscu, Stephen S. Rich

## Abstract

Type 1 diabetes is a chronic autoimmune disease caused by destruction of the pancreatic β-cells. Genome-wide association (GWAS) and fine mapping studies associated with type 1 diabetes are mainly in European ancestry populations. We conducted a multi-ancestry GWAS to identify single nucleotide polymorphisms (SNPs) and HLA alleles associated with type 1 diabetes risk and age at onset.

The Type 1 Diabetes Genetics Consortium (T1DGC) samples were genotyped using the Illumina CoreExome BeadChip array, and included families of largely European ancestry (EUR, N=3,222), unrelated individuals of African ancestry (AFR, N=891) and admixed (primarily Hispanic/Latino) ancestry (AMR, N=308). The four-digit multi-ancestry HLA reference panel (HLA-TAPAS) was used to impute HLA alleles. Logistic mixed models were used for analysis of genetic data with type 1 diabetes risk, while frailty models were used for analysis of age at onset.

Seven loci were associated with type 1 diabetes at genome-wide significance (P<5.0A×10^-8^) in the meta-analysis of the three ancestry groups: *PTPN22*, *HLA-DQA1*, *IL2RA*, *RNLS*, *INS*, *IKZF4-RPS26-ERBB3*, and *SH2B3*. Four loci were associated with age at onset (*PTPN22*, *HLA-DQB1*, *INS*, and *ERBB3*). AFR and AMR meta-analysis revealed that the *NRP1* locus was associated with both risk and age at onset; however, variants within *NRP1* were not significantly associated with risk in those of EUR ancestry. In contrast, the *PTPN22* variant rs2476601 (R620W) was significantly associated with risk only in EUR ancestry individuals. The HLA haplotype most significantly associated with risk in AFR and AMR ancestry was *HLA-DRB1*03:01-DQA1*05:01-DQB1*02:01* differed from the *HLA-DRB1*04:01-DQA1*03:01-DQB1*03:02* haplotype associated with risk in EUR ancestry. The *HLA-DRB1*08:02-DQA1*04:01-DQB1*04:02* haplotype was “protective” in AMR ancestry while *HLA-DRB1*08:01-DQA1*04:01-DQB1*04:02* (differing only at DRB1) was “risk” in EUR ancestry. The multi-ancestry GWAS enabled identification of ancestry-specific genetic variants, HLA alleles and haplotypes that are associated with type 1 diabetes risk and age at onset.

**Author Summary:** Twin and family studies have shown that genetics play important role in development of type 1 diabetes in childhood. The majority of genetics studies of type 1 diabetes have been performed in populations of European ancestry. Here, we examine the influence of common genetic variants and HLA haplotypes in diverse ancestry samples of type 1 diabetes and earlier age at disease onset. We show that *NRP1* region exhibits stronger association with type 1 diabetes risk and age at onset in non-European populations and *PTPN22* differentiates the risk of type 1 diabetes between individuals of European and non-European ancestries. In addition, we identify protective DR-DQ HLA haplotype in Admixed individuals and DR-DQ HLA haplotype associated with increased risk for type 1 diabetes in European ancestry population. These findings could help improve identification of individuals at high genetic risk of type 1 diabetes and provide opportunities for delay or prevention of type 1 diabetes.

## Introduction

Type 1 diabetes is a common autoimmune disease in which the destruction of pancreatic β-cells results in the eventual inability of the body to produce insulin [1–4]. Without insulin, there is accumulation of glucose in the bloodstream and an inability for glucose to enter cells for production of energy. Progression of glucose accumulation leads to blood vessel and organ damage from dehydration, conversion of tissue to ketones for alternative energy sources, and life-threatening diabetic ketoacidosis [5–8]. Thus, external sources of insulin are necessary for survival.

There are many risk factors associated with the development of type 1 diabetes, including both genetic and generally unknown environmental factors [9–11]. As a disease of autoimmunity, type 1 diabetes has been characterized by three specific stages [12]: Stage 1 represents the transition from normal glucose homeostasis in an individual with variable genetic and other risk factors to production of multiple islet autoantibodies but with glucose levels in the normal range; Stage 2 includes individuals who have multiple islet autoantibodies but with glucose levels exceeding normal range (e.g., fasting plasma glucose ≥ 100 mg/dL or ≥ 5.6 mmol/L); with Stage 3 representing clinically diagnosed type 1 diabetes. Each of these stages of type 1 diabetes may have overlapping, as well as distinct, genetic and non-genetic risk factors. Although type 1 diabetes has historically been thought to be a disease of childhood (known previously as juvenile-onset diabetes mellitus) and of Northern European ancestry, it occurs in individuals of all ages and ancestry groups [13–17].

The genetic basis of type 1 diabetes is well-established. Twin and family studies have estimated the genetic contribution to type 1 diabetes in childhood as roughly 50%. Early studies focused on the HLA region [18–20] and identified the contribution of the genes, alleles and haplotypes of the human Major Histocompatibility Complex (MHC) that have large effects on risk, specifically the HLA class I genes (*-A*, *-B*, and *-C*) and the HLA class II genes (*-DRB1*, *-DQA1*, *-DQB1*, *-DPA1*, *-DPB1*) [19–22]. Subsequent studies utilized candidate gene approaches (related to the immune response) with few (typically functional) genetic variants in small numbers of cases, controls, and families. The insulin gene (*INS*) variable number of tandem repeat (VNTR) polymorphism was a candidate gene identified through its impact on insulin metabolism and was implicated in risk of type 1 diabetes [23] and additional loci containing variants discovered with large effect that were in coding or promoter regions in candidate genes (e.g., *PTPN22* [24] and *CTLA4* [25]).

With the development of rapid genotyping technology and ability to assemble large numbers of cases with type 1 diabetes and controls, genome-wide association studies (GWAS) became a common tool to detect genetic variants associated with disease and risk factors [26]. Initial GWAS studies identified loci that had statistical power for the detection of common variants (minor allele frequency (MAF) > 0.05) with large effect (OR > 1.5) [26], there was recognition that much of the genetic impact on common human disease would have much smaller effects, requiring significantly increased sample size. The Type 1 Diabetes Genetics Consortium (T1DGC) was formed to conduct genome-wide analyses in affected sib-pair families (linkage) and case-control collections (association) to individually increase sample size and to conduct meta-analyses in type 1 diabetes [27]. Following GWAS, additional genotyping for fine mapping required development of a custom array (ImmunoChip) to better interrogate regions of interest across the genome [28,29]. Currently, GWAS meta-analyses and fine mapping studies [29,30] have identified over 100 loci associated with risk of type 1 diabetes.

A major limitation in most human genetic studies has been a focus on populations of European ancestry [31–33]. Despite increasing recognition of the benefit of including diversity in discovery, scientific equity, and reducing health disparities from applications of genetic medicine, there has yet to be a GWAS in type 1 diabetes that includes large sample sizes in non-European ancestry populations. This absence of study is a major gap, with the increasing global incidence and prevalence of type 1 diabetes (SEARCH) [34–36]. In this report, we conduct a multi-ancestry GWAS meta-analysis in cases, controls and families with type 1 diabetes for discovery of genetic variants, detection of novel ancestry-specific HLA variants and amino acid residues that are associated with risk of type 1 diabetes as well as the age at onset of disease.

## Results

A total of 13,412 individuals and 430,930 variants were included in our analyses after quality control and sample/variant filtering (**Supplementary Table 1**, **Supplementary Figure 1**). Each participant was assigned to one of three genetic ancestry groups – African (AFR), Admixed (AMR) and European (EUR). Association analyses were conducted separately on individual case-control groups (409 AFR cases, 482 AFR controls, 153 AMR cases, 155 AMR controls, and 3,428 pseudo-cases and 3,428 pseudo-controls generated from affected sibpair families). Genotypes were imputed using the TOPMed multi-ancestry reference panel, following SNP quality control, we used 13,777,800 (AFR), 8,952,895 (AMR), and 8,500,361 (pseudo case-control) common/infrequent variants (MAF ≥ 0.01) for association analyses.

### Genome-wide association analysis of type 1 diabetes across diverse ancestry

Logistic mixed models were fit for each group. In meta-analysis of AFR, AMR and pseudo case-control datasets, there was no evidence of systematic bias (λ_GC_ = 1.01) after excluding SNPs in the MHC region. Seven known type 1 diabetes-associated loci were identified at genome-wide significance (P < 5.0 x 10^-8^): 1p13.2 (*PTPN22*), 6p21.32 (HLA*-DQA1*), 10p15.1 (*IL2RA*), 10q23.31 (*RNLS*), 11p15.5 (*INS*), 12q13.2 (*IKZF4-RPS26-ERBB3*), and 12q24.12 (*SH2B3*) (**Table 1**; **Supplementary Figure 2A, Supplementary Figure 2B, Supplementary Figure 3A-3F**). The *INS* locus SNP rs689 exhibited the strongest association among non-HLA region SNPs (OR = 1.81, P = 2.34 x 10^-45^). Another non-HLA region lead variant, rs6679677 in the *PTPN22* locus, was in a high linkage disequilibrium (LD) (r^2^ ∼ 0.96) with the known rs2476601 (R620W) SNP, residing in the coding sequence of *PTPN22* gene [24,37,38]. However, the risk allele frequency of rs2476601 (in perfect LD with rs6679677 in EUR population) differs across ancestry groups and only the EUR group showed robust evidence of association with type 1 diabetes with comparable effect sizes across ancestries. The 12q13.2 region is complex with multiple potential candidate genes associated with type 1 diabetes, including *IKZF4*, *RPS26*, and *ERBB3*. Previously, a EUR ancestry GWAS identified *ERBB3* as a putative candidate gene [27], while fine-mapping studies supported the *IKZF4-RPS26* region [28,29]. In our diverse ancestry meta-analysis, rs7302200 was identified as the lead variant 12q13.2 (OR = 1.31, P = 7.74 x 10^-13^), residing between *RPS26* and *ERBB3*. Another type 1 diabetes-associated region on chromosome 12 contains the *SH2B3* locus, with rs597808 being the most significant variant (OR = 1.27, P = 4.82 x 10^-11^) and in high linkage disequilibrium (r^2^ > 0.96) with previously identified variants in this region (rs653178 [28] and rs7310615 [29]).

**Table 1.**
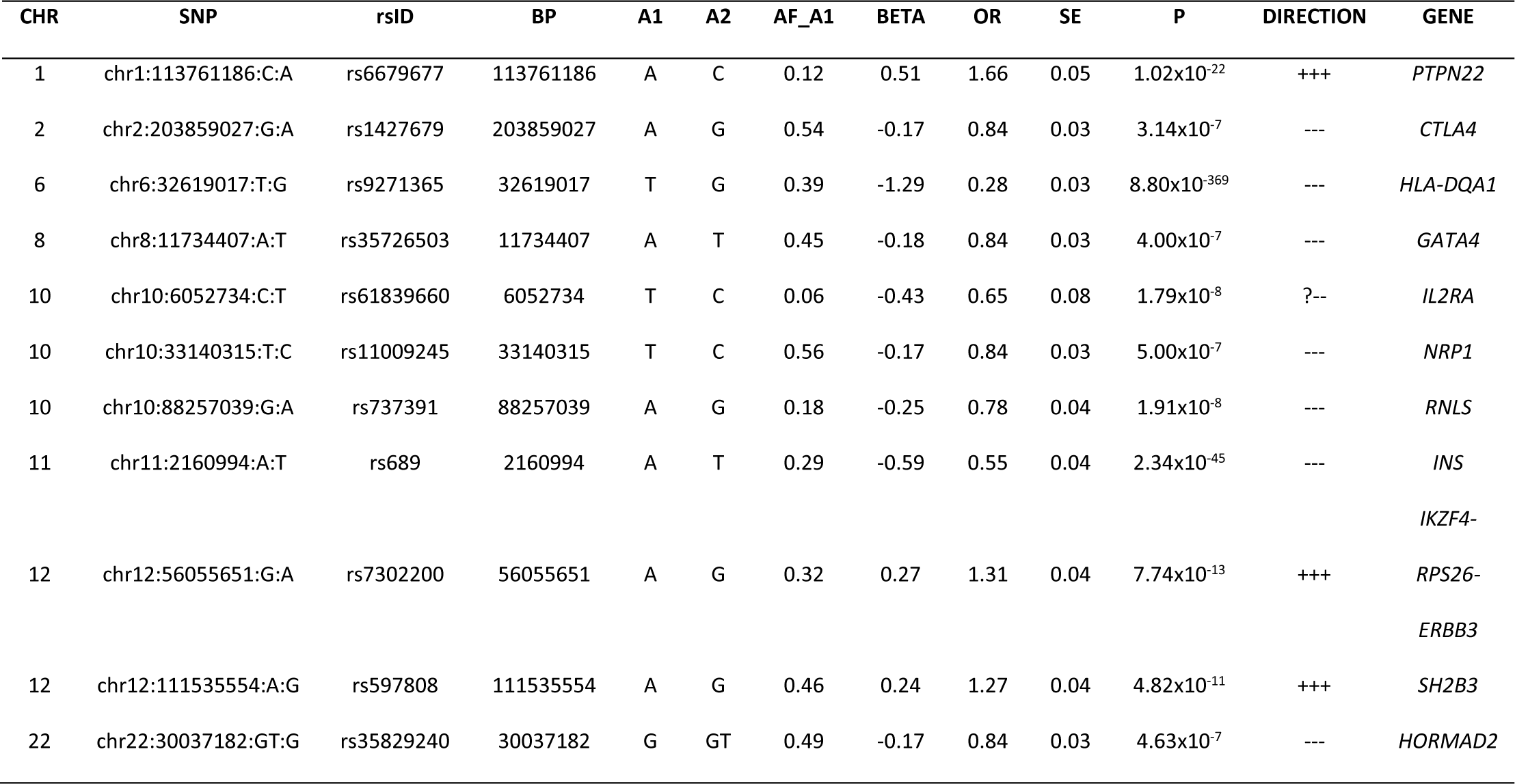
Lead variants associated with type 1 diabetes risk (SAIGE).

Four regions reached suggestive levels of genome-wide significance (P < 5.0 x 10^-7^): 2q33.2 (*CTLA4*), 8p23.1 (*GATA4*), 10p11.22 (*NRP1*) and 22q12.2 (*HORMAD2*). Evidence of association for the *NRP1* locus was strongest in the AFR and AMR populations where it reached the genome-wide significance (rs722988, OR_AFR_AMR_ = 1.61, P_AFR_AMR_ = 1.10 x 10^-8^) (**Supplementary Figure 4A**), while there was minimal evidence of association in the EUR population (OR_EUR_ = 1.11, P_EUR_ = 0.005; P_diff_ = 0.04). Previously, a large fine-mapping study identified the *NRP1* locus (rs722988, OR = 1.11, P = 3.21 x 10^-15^) as significantly associated with type 1 diabetes risk [29]. Results by ancestry can be found in **Supplementary Table 2**.

### Genome-wide association analysis of type 1 diabetes age at onset across diverse ancestry

AFR (N = 891), and AMR (N = 308) individuals with self-reported age at onset of type 1 diabetes and age-at-enrollment in the study for controls were used in the analyses. In the pseudo case-control dataset (N = 6,840) self-reported age at onset of type 1 diabetes, for both cases and pseudo-controls, was used for analysis. Censored time-to-event models were used for analysis for each group. There was no evidence of systematic bias (λ_GC_ = 1.01) after excluding SNPs in the MHC region. Meta-analysis of the three case-control groups for age at onset revealed four regions that attained genome-wide significance, all regions are established type 1 diabetes risk loci: 1p13.2 (*PTPN22*), 6p21.32 (HLA*-DQB1*), 11p15.5 (*INS*), and 12q13.2 (*ERBB3*) (**Table 2; Supplementary Figure 2C, Supplementary Figure 2D, Supplementary Figure 3G-3I**). Among the non-HLA region SNPs, rs689 in the *INS* locus had the strongest association with age at onset (OR = 1.45, P = 2.81 x 10^-28^), similar to its association with type 1 diabetes risk (OR = 1.81, P = 2.34 x 10^-45^). Although the association with age at onset was weaker, the trend was consistent with disease risk. Meta-analysis revealed that rs11171747, located near the *ERBB3* gene in the 12q13.2 region, was the most significantly associated SNP with age at onset (OR = 1.17, P = 1.20 x 10^-8^). This finding suggests that different (statistically independent) SNPs in the *IKZF4*-*RPS26*-*ERBB3* locus may be associated with type 1 diabetes risk and age at onset.

**Table 2.** Lead variants associated with type 1 diabetes age at onset (GATE).

| CHR | SNP | rsID | BP | A1 | A2 | AF_A1 | BETA | OR | SE | P | DIRECTION | GENE |
| --- | --- | --- | --- | --- | --- | --- | --- | --- | --- | --- | --- | --- |
| 1 | chr1:113761186:C:A | rs6679677 | 113761186 | A | C | 0.12 | 0.27 | 1.31 | 0.04 | $4.34 \times 10^{-11}$ | +++ | *PTPN22* |
| 6 | chr6:32658525:T:G | rs9273364 | 32658525 | T | G | 0.50 | -0.94 | 0.39 | 0.03 | $2.12 \times 10^{-301}$ | --- | *HLA-DQB1* |
| 10 | chr10:88287593:T:G | rs2018705 | 88287593 | T | G | 0.77 | 0.16 | 1.18 | 0.03 | $1.11 \times 10^{-7}$ | +++ | *RNLS* |
| 11 | chr11:2160994:A:T | rs689 | 2160994 | A | T | 0.30 | -0.37 | 0.69 | 0.03 | $2.81 \times 10^{-28}$ | --- | *INS* |
| 12 | chr12:56124624:T:G | rs11171747 | 56124624 | T | G | 0.64 | 0.16 | 1.17 | 0.03 | $1.20 \times 10^{-8}$ | +++ | *ERBB3* |
| 12 | chr12:111270654:A:C | rs1265564 | 111270654 | A | C | 0.58 | -0.15 | 0.86 | 0.03 | $1.38 \times 10^{-7}$ | --- | *SH2B3* |
| 18 | chr18:12857336:G:A | rs534911 | 12857336 | A | G | 0.74 | -0.16 | 0.85 | 0.03 | $2.95 \times 10^{-7}$ | --- | *PTPN2* |

Two regions associated with type 1 diabetes also reached suggestive evidence of association with age at onset: 10q23.31 (*RNLS*), 12q24.12 (*SH2B3*). The most strongly associated SNP with age at onset of type 1 diabetes at the *SH2B3* locus was rs1265564 (OR = 1.16, P = 1.38 x 10^-7^), which is in moderate linkage disequilibrium (r^2^ ∼ 0.60) with rs597808, the variant identified as associated with type 1 diabetes. The most strongly associated SNP in *RNLS* (rs2018705, OR = 1.18, P = 1.11 x 10^-7^) was similar in direction of effect for type 1 diabetes risk. One region 18p11.21 (*PTPN2*), reached suggestive significance for association with type 1 diabetes age at onset but not risk, although this region has been established previously [39,40]. In the *NRP1* locus, the same variant associated with type 1 diabetes risk and age at onset (rs722988, OR = 1.41, P = 2.41 x 10^-8^) reached the genome-wide significance only in the meta-analysis of AFR and AMR ancestry samples (**Supplementary Figure 4B**).

### HLA region class II gene and haplotype analysis in type 1 diabetes

The association analyses of type 1 diabetes with the HLA region included AFR ancestry (409 cases and 482 controls), AMR ancestry (153 cases, 155 controls), and EUR ancestry (2,970 pseudo-cases, 2,970 pseudo-controls). After imputation, there were 20,329 variants for (AFR), 20,376 variants (AMR), and 20,279 variants (EUR) in the HLA region; from these data classical HLA alleles and HLA gene amino acid sequences were imputed as described (Methods). The HLA*-DQA1*03:01* allele was the most significantly associated for both type 1 diabetes risk and age at onset in AFR and AMR groups. In contrast, the HLA*-DQB1*03:02* allele was most associated with both type 1 diabetes risk and age at onset in EUR ancestry subjects. The lead association with type 1 diabetes risk (**Figure 1**) and age at onset (**Figure 2**) was with the established HLA-DQB1 amino acid position 57 in all three-ancestry groups.

**Figure 1.**
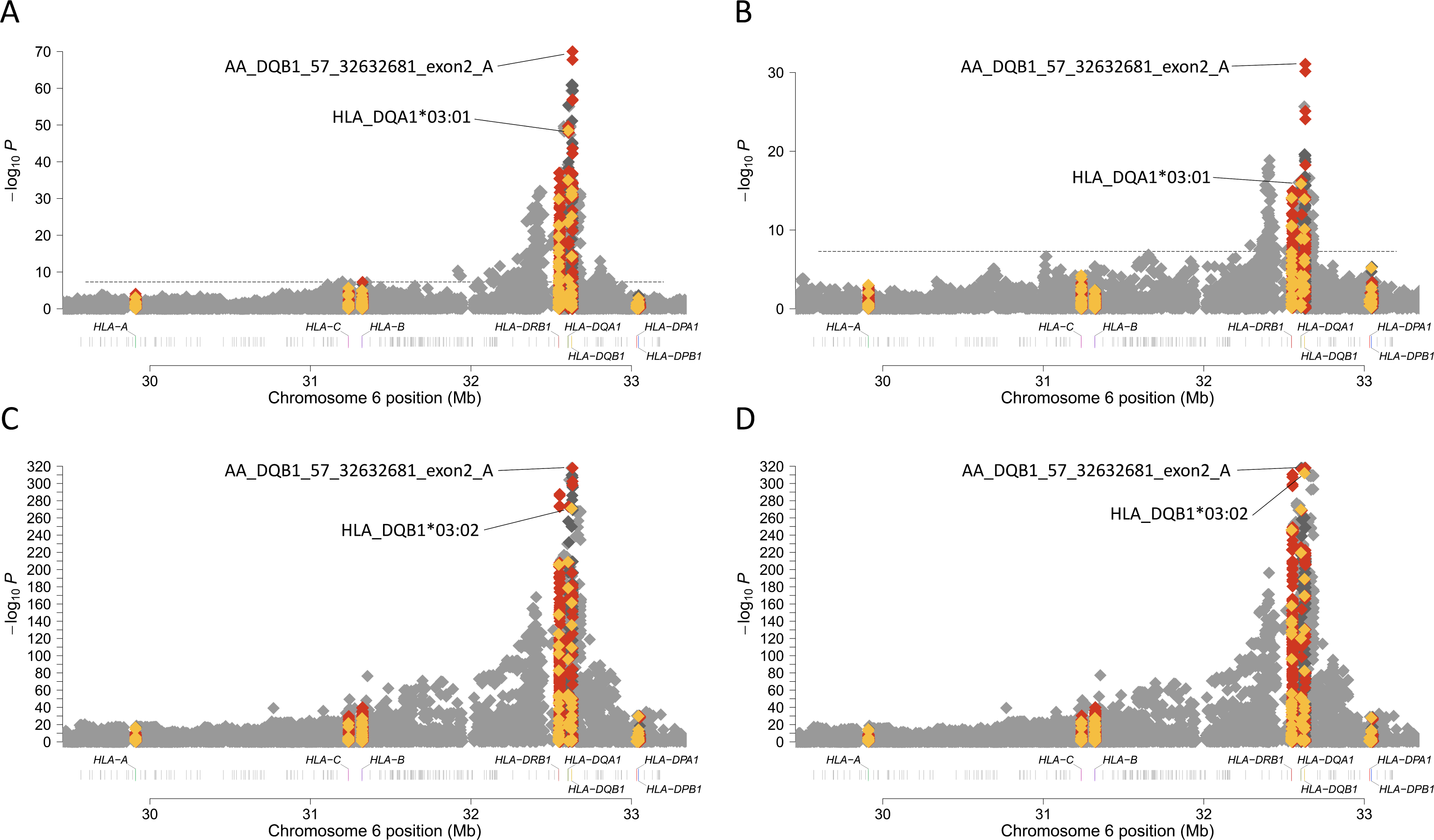
HLA alleles and amino acids associated with type 1 diabetes risk in AFR (**A**), AMR (**B**), EUR (**C**) and meta-analysis (**D**). HLA class I and HLA class II genes are labeled on the x-axis. The y-axis represents – log_10_(P-value). The horizontal dashed line represents the threshold for genome-wide significance. SNPs are represented by grey diamonds, HLA alleles by yellow diamonds, and HLA amino acids by red diamonds. (**A**) Type 1 diabetes risk associations within the HLA region in AFR ancestry individuals. (**B**) Type 1 diabetes risk associations within the HLA region in AMR ancestry individuals. (**C**) Type 1 diabetes risk associations within the HLA region in EUR ancestry individuals. (**D**) Type 1 diabetes risk associations within the HLA region in meta-analysis of AFR, AMR and EUR ancestry individuals.

**Figure 2.**
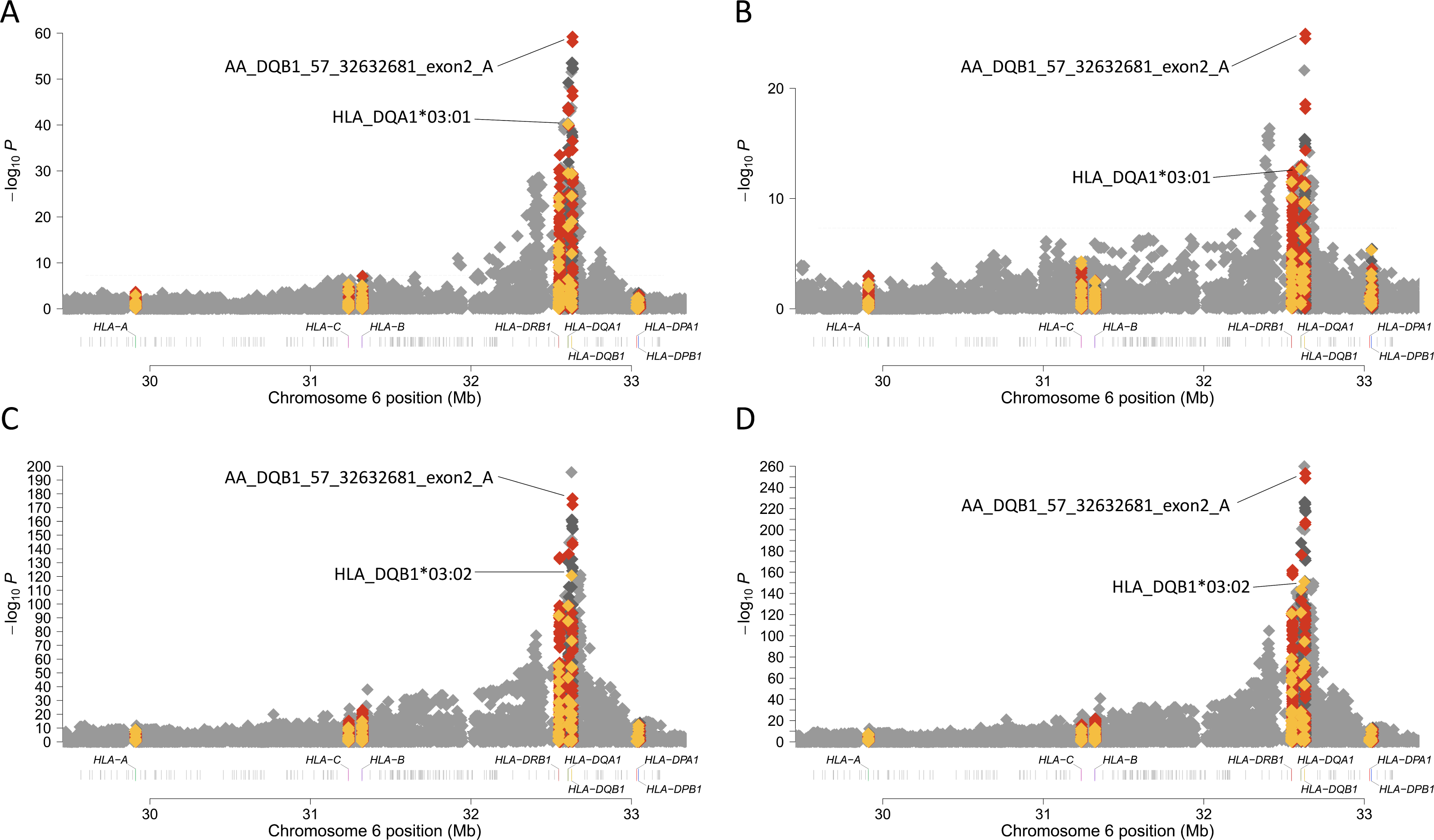
HLA alleles and amino acids associated with type 1 diabetes age at onset in AFR (**A**), AMR (**B**), EUR (**C**) and meta-analysis (**D**). HLA class I and HLA class II genes are labeled on the x-axis. The y-axis represents –log_10_(P-value). The horizontal dashed line represents the threshold for genome-wide significance. SNPs are represented by grey diamonds, HLA alleles by yellow diamonds, and HLA amino acids by red diamonds. (**A**) Type 1 diabetes age at onset associations within the HLA region in AFR ancestry individuals. (**B**) Type 1 diabetes age at onset associations within the HLA region in AMR ancestry individuals. (**C**) Type 1 diabetes age at onset associations within the HLA region in EUR ancestry individuals. (**D**) Type 1 diabetes age at onset associations within the HLA region in meta-analysis of AFR, AMR and EUR ancestry individuals.

It has been documented that the combination of specific *DRB1*, *DQA1* and *DQB1* alleles have been associated with the risk of type 1 diabetes [41,42]. To identify the ancestry-specific risk alleles of DR-DQ haplotypes, three locus HLA class II haplotypes were analyzed. The most significantly associated haplotype with type 1 diabetes in AFR and AMR (**Table 3**) was HLA-*DRB1*\*03:01-HLA-*DQA1*\*05:01-HLA-*DQB1*\*02:01 (OR_AFR_ = 4.23, P_AFR_ = 1.9 x 10^-22^; OR_AMR_ = 6.95, P_AMR_ = 2.6 x 10^-10^). In EUR pseudo case-controls (**Table 3**), the HLA-*DRB1*\*04:01-HLA-*DQA1*\*03:01-HLA-*DQB1*\*03:02 haplotype (OR_EUR_ = 6.66, P_EUR_ = 4.5 x 10^-207^) was the most significantly associated with type 1 diabetes.

**Table 3.** Association of MHC class II haplotypes with type 1 diabetes in pseudo case-control European, African and Admixed ancestry individuals.

| DRB1 | DQA1 | DQB1 | EUR |  |  |  | AFR |  |  |  | AMR |  |  |  |
| --- | --- | --- | --- | --- | --- | --- | --- | --- | --- | --- | --- | --- | --- | --- |
|  |  |  | control | case | OR* | P | control | case | OR* | P | control | case | OR* | P |
|  |  |  | AF | AF |  |  | AF | AF |  |  | AF | AF |  |  |
| 01:01 | 01:01 | 05:01 | 0.097 | 0.064 | 0.63 | $2.3 \times 10^{-10}$ | 0.026 | 0.032 | 1.16 | $6.2 \times 10^{-1}$ | . | . | . | . |
| 01:02 | 01:01 | 05:01 | 0.014 | 0.008 | 0.54 | $1.1 \times 10^{-3}$ | 0.031 | 0.030 | 0.99 | $9.8 \times 10^{-1}$ | . | . | . | . |
| 01:03 | 01:01 | 05:01 | 0.009 | 0.004 | 0.44 | $1.2 \times 10^{-3}$ | . | . | . | . | . | . | . | . |
| 03:01 | 05:01 | 02:01 | 0.152 | 0.332 | 3.10 | $5.0 \times 10^{-117}$ | 0.069 | 0.245 | 4.23 | $1.9 \times 10^{-22}$ | 0.048 | 0.233 | 6.95 | $2.6 \times 10^{-10}$ |
| 03:02 | 04:01 | 04:02 | . | . | . | . | 0.056 | 0.009 | 0.17 | $1.1 \times 10^{-7}$ | . | . | . | . |
| 04:01 | 03:01 | 03:01 | 0.042 | 0.020 | 0.48 | $7.8 \times 10^{-11}$ | . | . | . | . | . | . | . | . |
| 04:01 | 03:01 | 03:02 | 0.062 | 0.261 | 6.66 | $4.5 \times 10^{-207}$ | 0.016 | 0.088 | 6.17 | $6.3 \times 10^{-12}$ | . | . | . | . |
| 04:02 | 03:01 | 03:02 | 0.017 | 0.042 | 2.65 | $4.5 \times 10^{-15}$ | . | . | . | . | 0.008 | 0.107 | 12.88 | $8.8 \times 10^{-7}$ |
| 04:03 | 03:01 | 03:02 | 0.006 | 0.003 | 0.51 | $2.1 \times 10^{-2}$ | . | . | . | . | . | . | . | . |
| 04:04 | 03:01 | 03:02 | 0.036 | 0.055 | 1.60 | $5.8 \times 10^{-7}$ | . | . | . | . | 0.072 | 0.088 | 1.31 | $4.1 \times 10^{-1}$ |
| 04:05 | 03:01 | 02:01 | 0.002 | 0.007 | 3.39 | $1.1 \times 10^{-4}$ | . | . | . | . | . | . | . | . |
| 04:05 | 03:01 | 03:02 | 0.007 | 0.026 | 4.21 | $1.1 \times 10^{-16}$ | 0.009 | 0.084 | 9.95 | $1.4 \times 10^{-14}$ | . | . | . | . |
| 04:07 | 03:01 | 03:01 | 0.008 | 0.001 | 0.06 | $1.1 \times 10^{-11}$ | . | . | . | . | . | . | . | . |
| 04:07 | 03:01 | 03:02 | . | . | . | . | . | . | . | . | 0.088 | 0.099 | 1.20 | $5.5 \times 10^{-1}$ |
| 07:01 | 02:01 | 02:01 | 0.100 | 0.039 | 0.37 | $1.9 \times 10^{-36}$ | 0.075 | 0.057 | 0.73 | $1.4 \times 10^{-1}$ | 0.076 | 0.046 | 0.50 | $8.4 \times 10^{-2}$ |
| 07:01 | 02:01 | 03:03 | 0.027 | 0.002 | 0.07 | $8.3 \times 10^{-32}$ | . | . | . | . | . | . | . | . |

|  |  |  | EUR |  |  |  | AFR |  |  |  | AMR |  |  |  |
| --- | --- | --- | --- | --- | --- | --- | --- | --- | --- | --- | --- | --- | --- | --- |
| DRB1 | DQA1 | DQB1 | control | case | OR* | P | control | case | OR* | P | control | case | OR* | P |
|  |  |  | AF | AF |  |  | AF | AF |  |  | AF | AF |  |  |
| 07:01 | 03:01 | 02:01 | . | . | . | . | 0.008 | 0.049 | 6.68 | 8.2x10<sup>-8</sup> | . | . | . | . |
| 08:01 | 04:01 | 04:02 | 0.025 | 0.031 | 1.25 | 5.8x10<sup>-2</sup> | . | . | . | . | . | . | . | . |
| 08:02 | 04:01 | 04:02 | . | . | . | . | . | . | . | . | 0.120 | 0.031 | 0.25 | 3.4x10<sup>-4</sup> |
| 08:04 | 04:01 | 03:01 | . | . | . | . | 0.038 | 0.009 | 0.24 | 1.1x10<sup>-4</sup> | . | . | . | . |
| 09:01 | 03:01 | 02:01 | . | . | . | . | 0.037 | 0.102 | 2.99 | 1.0x10<sup>-7</sup> | . | . | . | . |
| 09:01 | 03:01 | 03:03 | 0.013 | 0.011 | 0.87 | 4.3x10<sup>-1</sup> | . | . | . | . | . | . | . | . |
| 10:01 | 01:01 | 05:01 | 0.008 | 0.002 | 0.22 | 1.2x10<sup>-6</sup> | . | . | . | . | . | . | . | . |
| 11:01 | 01:02 | 06:02 | . | . | . | . | 0.036 | 0.000 | 0.00† | 3.0x10<sup>-9</sup> | . | . | . | . |
| 11:01 | 05:01 | 03:01 | 0.056 | 0.009 | 0.16 | 4.2x10<sup>-47</sup> | 0.032 | 0.011 | 0.32 | 2.1x10<sup>-3</sup> | . | . | . | . |
| 11:02 | 05:01 | 03:01 | . | . | . | . | 0.052 | 0.009 | 0.17 | 3.1x10<sup>-7</sup> | . | . | . | . |
| 11:04 | 05:01 | 03:01 | 0.029 | 0.003 | 0.08 | 4.0x10<sup>-34</sup> | . | . | . | . | . | . | . | . |
| 12:01 | 01:01 | 05:01 | . | . | . | . | 0.032 | 0.011 | 0.33 | 3.1x10<sup>-3</sup> | . | . | . | . |
| 12:01 | 05:01 | 03:01 | 0.013 | 0.004 | 0.32 | 2.2x10<sup>-7</sup> | . | . | . | . | . | . | . | . |
| 13:01 | 01:03 | 06:03 | 0.057 | 0.012 | 0.19 | 3.7x10<sup>-42</sup> | . | . | . | . | . | . | . | . |
| 13:02 | 01:02 | 06:04 | 0.036 | 0.026 | 0.72 | 3.0x10<sup>-3</sup> | 0.014 | 0.024 | 1.72 | 1.5x10<sup>-1</sup> | . | . | . | . |
| 13:02 | 01:02 | 06:09 | . | . | . | . | 0.019 | 0.017 | 0.89 | 7.5x10<sup>-1</sup> | . | . | . | . |
| 13:03 | 05:01 | 03:01 | 0.010 | 0.002 | 0.18 | 3.0x10<sup>-9</sup> | . | . | . | . | . | . | . | . |
| 14:01 | 01:01 | 05:03 | 0.013 | 0.001 | 0.04 | 3.0x10<sup>-19</sup> | . | . | . | . | . | . | . | . |
| 15:01 | 01:02 | 06:02 | 0.107 | 0.004 | 0.03 | 6.2x10<sup>-151</sup> | . | . | . | . | . | . | . | . |
| 15:02 | 01:03 | 06:01 | 0.008 | 0.001 | 0.11 | 4.8x10<sup>-10</sup> | . | . | . | . | . | . | . | . |
| 15:03 | 01:02 | 06:02 | . | . | . | . | 0.123 | 0.015 | 0.11 | 1.1x10<sup>-18</sup> | . | . | . | . |
| 16:01 | 01:02 | 05:02 | 0.020 | 0.013 | 0.64 | 3.5x10<sup>-3</sup> | . | . | . | . | . | . | . | . |

\* OR adjusted
† test based on 0 cases and 31 controls

Using conditional analysis, we found seven AFR HLA haplotypes (**Supplementary Table 3)**, two AMR HLA haplotypes (**Supplementary Table 4)**, and nineteen EUR HLA haplotypes (**Supplementary Table 5**) independently associated with type 1 diabetes. Conditional analysis revealed that HLA-*DRB1*\*08:02-HLA-*DQA1*\*04:01-HLA-*DQB1*\*04:02 haplotype was protective (OR < 1) in AMR population (OR = 0.39, P = 2.9 x 10^-2^), while in EUR population HLA-*DRB1*\*08:01-HLA-*DQA1*\*04:01-HLA-*DQB1*\*04:02 was associated with increased risk for type 1 diabetes (OR = 1.81, P = 5.5 x 10^-5^).

### Enrichment of type 1 diabetes genes across autoimmune diseases (FUMA)

The coexistence of type 1 diabetes with other immune-mediated diseases has been extensively documented, thus we compared the enrichment of T1D annotated genes against GWAS-catalog reported genes in autoimmune diseases (**Figure 3**). We identified 13 autoimmune diseases that shared annotated genes, including type 1 diabetes. Excluding type 1 diabetes, the most significant over-representation of identified genes in autoimmune diseases was in Alopecia Areata (P = 3.62 x 10^-16^). The overlap between ‘T1D’ identified loci and ‘AA’ was driven, in part, by common susceptibility variants within *PTPN22*, *IL2RA*, *IKZF4*, *ERBB3* and *SH2B3* loci. The disease that had the largest number of overlapping genes with type 1 diabetes was Crohn’s disease (P = 1.54 x 10^-7^); however, the direction of effect for the associated variants were not highly concordant, thus decreasing the genetic similarity despite a large number of associated loci. In addition to their genetic similarity with type 1 diabetes across ancestries, most of the identified diseases have characteristic tissue autoantibodies (e.g., AA, JIA, and RA), as shown previously in EUR ancestry populations [28].

**Figure 3.**
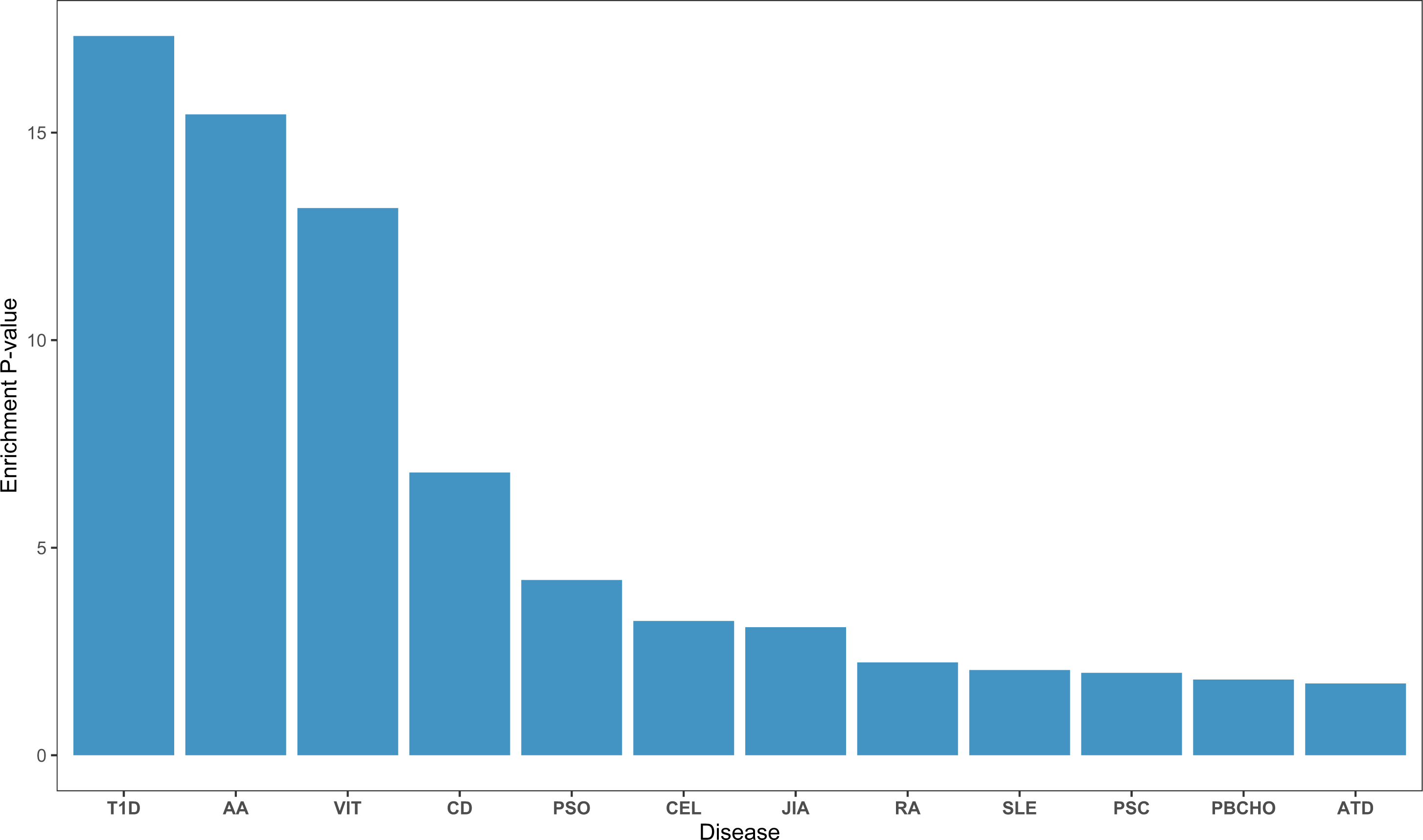
Enrichment of T1D identified genes in other autoimmune diseases. Each bar is labeled with the adjusted P-value using a multiple test correction (Benjamini–Hochberg). The MHC region (chr6: 25Mb-35Mb) was excluded from analysis. AA - Alopecia Areata, ATD - Autoimmune Thyroid Disease, CEL - Celiac Disease, CD - Crohn’s Disease, JIA - Juvenile Idiopathic Arthritis, PBCHO - Primary Biliary Cholangitis, PSC - Primary Sclerosing Cholangitis, PSO - Psoriasis (PSO), RA - Rheumatoid Arthritis, SLE - Systemic Lupus Erythematosus, T1D - Type 1 diabetes, VIT – Vitiligo.

## Discussion

In this multi-ancestry genome-wide meta-analysis of type 1 diabetes, we identified seven regions at genome-wide significance associated with risk and four regions at genome-wide significance associated with age at onset, with some regions exhibiting ancestry-specific effects. In the HLA region, we determined specific HLA class I and class II alleles and amino acids associated with type 1 diabetes risk and age at onset within and across ancestry, as well as ancestry-specific HLA haplotypes. We identified seven associated HLA haplotypes in African ancestry group, two in the admixed (Hispanic/Latino) ancestry group, and nineteen HLA haplotypes significantly associated with risk in the European ancestry group.

As expected, the strongest non-HLA regions associated with type 1 diabetes risk and age at onset were seen at the known *INS* and *PTPN22* loci. However, the most associated variant (rs2476601, C1858T, R620W, in complete linkage disequilibrium with rs6679677) in the *PTPN22* locus was only supported by European-ancestry. The association between *PTPN22* rs2476601 and type 1 diabetes was first documented in case-control study of non-Hispanic white individuals from North America and Sardinia [24]. Subsequent family and case-control studies in numerous European populations [37,43–46] confirmed its association with type 1 diabetes in this locus, showing high frequencies of rs2476601 in northern European populations and decreased frequencies in southern European and Sardinian populations [47]. The *PTPN22* rs2476601 variant has been shown to be rare in African and Asian populations [47–49], supporting our results of reduced association in non-EUR ancestry individuals. In addition, it has been shown that the *PTPN22* rs2476601 SNP is associated with earlier age at onset of type 1 diabetes in the European ancestry population [50].

We identified an association of the *NRP1* locus with type 1 diabetes risk and age at onset in non-European ancestry (African and admixed ancestry groups) but not in the European ancestry group. This locus has been identified in a large fine-mapping study involving diverse ancestry individuals [29] and genome-wide association study of a large European cohort [30]. Together, the *NRP1* locus may represent a region more associated with type 1 diabetes risk in non-European ancestry populations, as the effect sizes are stronger in the African and admixed groups; however, the direction of effect is the same in all ancestry groups. In addition, the strongest associated variant in the *NRP1* locus was associated with earlier age at onset. Previously, two SNPs located in intron 9 of *NRP1* were shown to be associated with type 1 diabetes [51], with the strength of association stronger in children with onset before age 10 years and/or in children who had a parent with type 1 diabetes. In addition, an *NRP1* isoform in pancreatic islets has been shown to be associated with a very young age at onset of type 1 diabetes [52]. Cells containing the truncated version of neuropilin-1 protein (encoded by *NRP1*) are devoid of insulin, resulting in the development of type 1 diabetes at a very early age. In those with onset of type 1 diabetes before age 4 years, the frequency of the minor allele (T) of the *NRP1* intron 9 variant (rs2070303) is increased when compared with subjects having an older age at onset.

Due to the importance of the HLA region in type 1 diabetes and limited data in non-European ancestries, we determined ancestry-specific haplotypes in collected cohorts. We identified the HLA-*DRB1*\*03:01-HLA-*DQA1*\*05:01-HLA-*DQB1*\*02:01 haplotype as the most significantly associated with type 1 diabetes in non-European ancestry individuals (African and Hispanic/Latino). A fine-mapping study of 3,949 African samples revealed the same haplotype as the strongest association [49]. In European ancestry subjects, the most significantly associated haplotype was HLA-*DRB1*\*04:01-HLA-*DQA1*\*03:01-HLA-*DQB1*\*03:02, consistent with previous results [53]. Comparisons across ancestries implicated differences in risk based upon HLA-*DRB1* gene, such as HLA-*DRB1*\*08:02-HLA-*DQA1*\*04:01-HLA-*DQB1*\*04:02 as a protective haplotype in the admixed population, with HLA-*DRB1*\*08:01-HLA-*DQA1*\*04:01-HLA-*DQB1*\*04:02 representing a susceptible haplotype in the European population.

This study has several strengths, including use of currently large sample sizes of under-represented populations (African and admixed ancestry), the genome-wide coverage of variants, and imputation to increase SNP density. In addition, the detailed interrogation of HLA region provides novel information on risk associations not only of HLA alleles but also amino acid residues and HLA haplotypes. There are, however, some limitations of the study, including the small number of samples compared with other genomic studies in European ancestry (despite having large non-EUR ancestry data). The relatively small number of samples in total may explain the number of type 1 diabetes-associated loci and failure to identify new regions that would likely have small effect sizes. Despite these limitations, it is important to recognize potential ancestry-specific effects on risk of type 1 diabetes, thereby better defining the genetic landscape of type 1 diabetes risk in non-EUR populations, particularly in the HLA region as it represents a major risk locus for type 1 diabetes.

Our multi-ancestry GWAS analysis enabled discovery of genetic variants, novel ancestry-specific HLA variants and amino acid residues that are associated with risk of type 1 diabetes and age at onset of disease. The inclusion of subjects with diverse genetic ancestry revealed that *NRP1* region exhibits stronger effect on type 1 diabetes risk and age at onset in non-EUR populations. The association of *NRP1* locus and specific HLA haplotypes, in addition to *PTPN22* differentiate the risk of type 1 diabetes between individuals of European and non-European ancestries. These results suggest that further increasing samples diversity can provide better understanding of genetic risk factors contributing to type 1 diabetes in non-EUR populations. This could help improve population-specific genetic risk scores application to identify individuals at high genetic risk of type 1 diabetes and provide opportunities for islet autoantibody screen for eligibility into early intervention trials (e.g., teplizumab) to delay or prevent disease onset.

## Methods

### Participants

We obtained DNA samples from 13,412 subjects recruited by the T1DGC (**Supplementary Table 1**). The study population consisted of 12,213 individuals from affected sib-pair and trio families. The majority of individuals from families were of European (EUR) ancestry (10,501). The case-control series consisted of 891 unrelated individuals of AFR ancestry (409 type 1 diabetes cases, 482 controls) and 308 individuals of AMR ancestry (153 type 1 diabetes cases, 155 controls).

### Genotyping and quality control

All samples were genotyped according to the manufacturer’s protocol using the Illumina Infinium CoreExome BeadChip in the Genome Sciences Laboratory at the University of Virginia. Raw genotyped data were subjected to SNP-level and sample-level quality control (**Supplementary Figure 1**) using KING software (version 2.2.8 [54]). For sample level quality control, we utilized chromosome X heterozygosity and chromosome Y absence to identify DNA samples that had discordant results between genetic sex and self-reported sex, these samples were treated as errors in processing and were removed. Additionally, we removed samples with a genotype call rate <95% and evidence of Mendelian inconsistences (MI, for families). Pedigree sample relationships were updated using KING software [54]. From samples passing initial quality control metrics, the following filters were applied for variants: removal of monomorphic SNPs, removal of SNPs with call rates <95%, removal of SNPs with Mendelian inconsistencies in >1% of the parent–offspring pairs and trios, removal of SNPs with low call rate in rare variants, and removal of SNPs significantly deviating from Hardy-Weinberg Equilibrium (P < 1.0 x 10^-6^ [P < 1.0 x 10^-20^ for MHC region]). Variant quality control included removal of SNPs that were not mapped uniquely to the genome (e.g., exm244817). Additional sample quality control included removal of family members with type 1 diabetes or case samples with age at onset of zero, and removal of family samples with onset of diabetes greater than 32 years and with at least one affected parent (suggestive of misdiagnosis of type 1 diabetes with maturity-onset diabetes of the young, MODY). All DNA samples were collected after approval from relevant institutional research ethics committees and appropriate informed consent was obtained from all subjects and families.

### Generation of a pseudo case-control sample

In the T1DGC, the family collection was ascertained specifically to include affected sib-pairs [55] followed by the collection of unrelated individuals with type 1 diabetes and controls. As the analytic method (logistic mixed models) is not robust to the targeted ascertainment of affected sib-pair families, a series of pseudo-cases and pseudo-controls was generated for application of the logistic mixed and frailty models (SAIGE and GATE). Within each family and for each SNP, the alleles transmitted from each parent to an affected child constituted the “pseudo case”; similarly, alleles not transmitted from each parent to an affected child constituted the “pseudo control”. From EUR affected sib-pair families, 3,428 pseudo-cases and 3,428 pseudo-controls were constructed; summary statistics from the pseudo case-control analysis was used in meta-analysis with the AFR and AMR case-control results.

### Population stratification

To infer genetic ancestry, we used multi-dimensional scaling (MDS) analysis implemented in KING [54]. We utilized the 1000 Genomes phase-3 reference panel to define the principal components and projected T1DGC subjects to this space to define subjects with EUR, AFR and AMR ancestry. Ancestry-specific principal components (PCs) were generated with principal component analysis (PCA) in control individuals, using SNPs selected by excluding the MHC region, removing SNPs with MAF > 0.05, and pruning for linkage disequilibrium (r^2^ < 0.5 in 50-kb windows). Genotypes of cases were projected onto control samples using PLINK v1.9 [56] to “match” for ancestry and minimize stratification effects in case-control analyses. A total of six subjects were removed that represented “outliers” from the PCA projection of type 1 diabetes cases onto controls.

### Imputation

Genotypes from the Illumina CoreExome BeadChip were imputed to the Trans-Omics for Precision Medicine (TOPMed) reference panel [57] on the TOPMed Imputation Server housed on the NHLBI BioData Catalyst server (https://imputation.biodatacatalyst.nhlbi.nih.gov/). For the GWAS data, a *minimac4* imputation accuracy of r^2^ > 0.3 was used as a variant filter for common and infrequent variants (MAF > 0.01), with r^2^ > 0.5 used as a filter for rare variants (MAF ≤ 0.01). For both common and rare variants, all SNPs were removed with Mendelian inconsistencies (MI) in at least 10% of trio families or parent– offspring (PO) pairs.

HLA imputation was conducted using the multi-ancestry HLA reference panel (HLA-TAPAS) [58] at the University of Michigan (https://imputationserver.sph.umich.edu/index.html). SNPs in the HLA region (28Mbp - 34Mbp) were used to impute HLA alleles (to four-digit accuracy) and amino acid residues. The HLA-TAPAS reference panel was generated using whole genome sequencing data from ∼20,000 samples from five global populations [58]. Classical alleles for HLA class I (HLA-A, -B, and -C) and HLA class II (HLA-DQA1, -DQB1, -DRB1, -DPA1, and -DPB1) genes were inferred from reads extracted from the extended MHC region by applying a population reference graph for the MHC region. Imputation accuracy was assessed by comparing the ‘gold standard’ sequence-based typing in individuals from the 1000 Genomes Project and the Japanese cohort to inferred HLA classical alleles. For the HLA imputation, variants with r^2^ > 0.5 and MAF > 0.005 were retained. Variants were removed with MI > 10% in trio families or in parent-offspring pairs.

### Statistical Analyses

Analysis of GWAS data for type 1 diabetes risk (binary trait) was conducted using SAIGE software that implemented a logistic mixed model (LMM) regression approach to control for type 1 error by accounting for unbalanced case-control ratio and sample relatedness. For type 1 diabetes age at onset, GATE software was used for GWAS based on a frailty model under similar unbalanced conditions.

#### Type 1 diabetes risk (SAIGE)

GWAS data passing quality control filters (MAF > 0.01 and minor allele count (MAC) ≥ 20) were analyzed for association with type 1 diabetes within three groups (pseudo case-control with majority of EUR ancestry [n=6,856], AFR [n=891], and AMR [n=308]). Logistic mixed model (LMM) regression implemented in SAIGE software [59] was used for analysis, which accounts for sample relatedness, adjusting for four principal components in AFR and AMR case-control groups and seven principal components in the pseudo case-control group. SAIGE controls for type I error rates even for unbalanced case-control ratios by incorporating a saddlepoint approximation (SPA) to improve estimation of the test statistic distribution at the extremes. Results of single-variant association tests were combined using a fixed-effects meta-analysis using METAL software [60]. Association results for selected loci were plotted using LocusZoom [61,62]. To determine the effect of population stratification in association analysis, the genomic inflation factor (λ_GC_) was estimated, defined by the ratio of median of the empirically observed distribution of the test statistic to the expected median. We performed conditional analysis on genome-wide significant (or suggestive) variants to identify loci with more than one variant independently associated with type 1 diabetes risk by including the most associated SNP in the LMM and identifying the subsequent (statistically significant) SNP, adding that SNP to the model until significance was no longer present.

#### Type 1 diabetes age at onset (GATE)

Association of GWAS variants passing filters (MAF > 0.01 and MAC ≥ 20) was evaluated with age at onset of type 1 diabetes by analysis of three groups (pseudo-case pseudo-control with majority of EUR ancestry, AFR, and AMR). We used the frailty mixed model regression implemented in GATE software [63], adjusting for four principal components in AFR and AMR case-control groups and seven principal components in the pseudo case-control group. The age for all individuals with and without type 1 diabetes was censored at 32 years old. Results of single-variant association tests were combined using a fixed-effects meta-analysis with METAL software.

#### HLA analysis

Association analyses of type 1 diabetes risk with imputed HLA alleles and amino acid residues (MAF > 0.005 and MAC ≥ 10) were conducted in 3 ancestry-specific groups (EUR [N = 5,940], AFR [N = 891], AMR [N = 381]) using SAIGE software. Association analyses of type 1 diabetes age at onset using GATE software employed the same approaches. Due to missing age information, fewer samples were used for frailty mixed model analysis in EUR ancestry (n=5,822). All HLA region association analyses were adjusted for four principal components in all three-ancestry groups.

To analyze association of HLA class II haplotypes with type 1 diabetes, all HLA DRB1-DQA1-DQB1 haplotypes were identified by creating a matrix of all possible combinations of alleles for each sample in each ancestry-specific group. For any given locus, individuals were excluded from the analysis if the total allele count for all possible alleles at that locus did not equal two. Common HLA alleles were defined as those that occur more than 30 times in a combined group of case and control subjects. For each ancestry group (EUR, AFR, AMR), association analyses of type 1 diabetes with HLA class II haplotypes (DRB1-DQA1-DQB1) were conducted on common haplotypes using logistic regression model and adjusting for four principal components as described previously [49]. Independent associations of HLA DRB1-DQA1-DRB1 haplotypes were assessed by conditioning on the most significant haplotype. The process was repeated until the consecutive haplotype failed to meet the significance threshold. For each ancestry group, the statistical significance was corrected for the total number of common haplotypes.

### Functional impact of detected variants

GWAS summary statistics were applied to the *SNP2GENE* and *GENE2FUNCTION* modules in FUMA v1.5.3 (Functional Mapping and Annotation, https://fuma.ctglab.nl). The *SNP2GENE* module was used to functionally annotate leading SNPs and the *GENE2FUNCTION* module was used to annotate genes and to identify genes that were enriched in pre-defined gene sets (e.g., reported genes from the GWAS-catalog). We focused on enrichment of genes in the autoimmune diseases from the GWAS-catalog gene set. Identified autoimmune diseases from GWAS-catalog gene set included: Alopecia Areata (AA), Autoimmune thyroid diseases (ATD), Celiac disease (CEL), Crohn’s disease (CD), Juvenile Idiopathic Arthritis (JIA), Primary Biliary cholangitis (PBC), Primary Sclerosing Cholangitis (PSC), Psoriasis (PSO), Rheumatoid Arthritis (RA), Systemic Lupus Erythematosus (SLE), Type 1 diabetes (T1D) and Vitiligo (VIT). Enrichment p-values were adjusted using a multiple test correction (Benjamini–Hochberg).

All statistical analyses and data visualization were performed using R version 4.1.1, unless otherwise stated. The HLA-TAPAS, Locus Zoom (http://locuszoom.org/) and R packages *ggplot2*, *qqman* and *RColorBrewer* were used for data visualization. For all conducted analyses genome-wide significance was based upon P < 5.0 x 10^-8^, while P < 5.0 x 10^-7^ was considered as suggestive evidence of association. The LD estimates were obtained from the LD pair tool (https://ldlink.nih.gov/) [64] with African (AFR), admixed (AMR) and European (EUR) cohorts selection.

## Data Availability

All summary statistics in the present study are being deposited into the T1D Knowledge Portal (https://t1d.hugeamp.org/), the database of Genotypes and Phenotypes (dbGaP, https://dbgap.ncbi.nlm.nih.gov/aa/wga.cgi?page=login) and the NHGRI-EBI GWAS Catalog (https://www.ebi.ac.uk/gwas/)

## Acknowledgments

The authors thank the investigators who assembled the collections of participants, obtained data and samples, and the participants who made the research possible, following groups and individuals who provided biological samples or data for this study. This research utilizes resources provided by the Type 1 Diabetes Genetics Consortium (T1DGC), a collaborative clinical study sponsored by the National Institute of Diabetes and Digestive and Kidney Diseases (NIDDK), National Institute of Allergy and Infectious Diseases, National Human Genome Research Institute, National Institute of Child Health and Human Development, and JDRF.

## Supporting information captions

**Supplementary Figure 1.** SNP and sample quality control workflow before imputation

**Supplementary Figure 2.** Loci associated with type 1 diabetes risk (**A, B**) and age at onset (**C, D**) in AFR, AMR and pseudo case-control meta-analysis. Each locus is labeled with the putative candidate gene. The y-axis represents –log(P values). The horizontal red line represents the threshold for genome-wide associations. (**A**) The strongest associations with type 1 diabetes risk (*HLA-DQA1*, *INS* and *PTPN22*). (**B**) Associations with type 1 diabetes risk after excluding HLA region (25 Mb – 35 Mb). (**C**) The strongest associations with age at onset of type 1 diabetes (*HLA-DQA1*, *INS* and *PTPN22*). (**D**) Associations with age at onset of type 1 diabetes after excluding HLA region (25 Mb – 35 Mb).

**Supplementary Figure 3.** LocusZoom plots of genome-wide significant loci associated with type 1 diabetes risk (**A-F**) and age at onset (**G-I**), excluding HLA region. The left y-axis represents –log(P values) and right y-axis determines recombination rate. The horizontal gray dashed line represents the threshold for genome-wide associations. The most significantly associated SNP is shown as purple diamond. For *PTPN22* locus (**A, G**), the purple diamond represents known coding region variant.

**Supplementary Figure 4.** Loci associated with type 1 diabetes risk (**A**) and age at onset (**B**) in non-EUR meta-analysis (AFR and AMR). Each locus is labeled with the putative candidate gene. The y-axis represents –log(P values). The horizontal red line represents the threshold for genome-wide associations. (**A**) Associations with type 1 diabetes risk in non-EUR meta-analysis. (**B**) Associations with age at onset of type 1 diabetes in non-EUR meta-analysis.

**Supplementary Table 1.** Number of genotyped individuals with phenotype data.

**Supplementary Table 2.** Type 1 diabetes risk within genetic ancestry.

**Supplementary Table 3.** Seven HLA class II haplotypes are independently associated with T1D in African-ancestry individuals. Odds ratios (OR) and P-values generated by multivariable logistic regression of T1D risk, including 4 principal components and all 7 alleles as independent variables.

**Supplementary Table 4.** Two HLA class II haplotypes are independently associated with T1D in Admixed-ancestry individuals. Odds ratios (OR) and P-values generated by multivariable logistic regression of T1D risk, including 4 principal components and 2 alleles as independent variables.

**Supplementary Table 5.** Nineteen HLA class II haplotypes are independently associated with T1D in European-ancestry individuals. Odds ratios (OR) and P-values generated by multivariable logistic regression of T1D risk, including 4 principal components and all 19 alleles as independent variables.

