## Supplemental Tables 1-5 for "A multi-ancestry genome-wide association study in type 1 diabetes"

**Supplementary Table 1.** Number of genotyped individuals with phenotype data

| Cohort | Total subjects | Cases (T1D) | Controls (unaffected) | Families | Females | Males | Mean age of controls enrollment (SD) | Mean age of T1D onset (SD) |
|---|---|---|---|---|---|---|---|---|
| AFR | 891 | 409 | 482 | . | 596 | 295 | 33.79 (13.10) | 9.33 (5.60) |
| AMR | 308 | 153 | 155 | . | 179 | 129 | 31.79 (11.40) | 8.31 (4.62) |
| Family ASP | 10833 | 5614 | 5180 | 2736 | 5540 | 5293 | 41.74 (16.37) | 11.01 (7.94) |
| Family trios | 1380 | 472 | 907 | 487 | 715 | 665 | 46.78 (11.67) | 9.62 (5.64) |
| Total | 13412 | 6648 | 6724 | 3223 | 7030 | 6382 | | |

* In families, 40 parents have missing phenotypes

**Supplementary Table 2.** Type 1 diabetes risk within genetic ancestry

| Meta-analysis (AFR, AMR, pseudo case-control) | | | | | | | | | | | AFR | | |
|---|---|---|---|---|---|---|---|---|---|---|---|---|---|
| **CHR** | **GENE** | **rsID** | **BP** | **A1** | **A2** | **AF_A1** | **BETA** | **OR** | **SE** | **P** | **AF_A1** | **BETA** | **OR** |
| 1 | *PTPN22* | rs6679677 | 113761186 | A | C | 0.12 | 0.51 | 1.66 | 0.05 | 1.02E-22 | 0.02 | 0.46 | 1.59 |
| 2 | *CTLA4* | rs1427679 | 203859027 | A | G | 0.54 | -0.17 | 0.84 | 0.03 | 3.14E-07 | 0.44 | -0.16 | 0.85 |
| 6 | *HLA-DQA1* | rs9271365 | 32619017 | T | G | 0.39 | -1.29 | 0.28 | 0.03 | 8.80E-369 | 0.45 | -1.18 | 0.31 |
| 8 | *GATA4* | rs35726503 | 11734407 | A | T | 0.45 | -0.18 | 0.84 | 0.03 | 4.00E-07 | 0.35 | -0.15 | 0.86 |
| 10 | *IL2RA* | rs61839660 | 6052734 | T | C | 0.06 | -0.43 | 0.65 | 0.08 | 1.79E-08 | . | . | . |
| 10 | *NRP1* | rs11009245 | 33140315 | T | C | 0.56 | -0.17 | 0.84 | 0.03 | 5.00E-07 | 0.37 | -0.23 | 0.80 |
| 10 | *RNLS* | rs737391 | 88257039 | A | G | 0.18 | -0.25 | 0.78 | 0.04 | 1.91E-08 | 0.09 | -0.28 | 0.76 |
| 11 | *INS* | rs689 | 2160994 | A | T | 0.29 | -0.59 | 0.55 | 0.04 | 2.34E-45 | 0.67 | -0.39 | 0.67 |
| 12 | *IKZF4-RPS26-ERBB3* | rs7302200 | 56055651 | A | G | 0.32 | 0.27 | 1.31 | 0.04 | 7.74E-13 | 0.07 | 0.18 | 1.19 |
| 12 | *SH2B3* | rs597808 | 111535554 | A | G | 0.46 | 0.24 | 1.27 | 0.04 | 4.82E-11 | 0.09 | 0.15 | 1.16 |
| 22 | *HORMAD2* | rs35829240 | 30037182 | G | GT | 0.49 | -0.17 | 0.84 | 0.03 | 4.63E-07 | 0.66 | -0.17 | 0.85 |
| **Meta-analysis (AFR, AMR)** | | | | | | | | | | | **AFR** | | |
| **CHR** | **GENE** | **rsID** | **BP** | **A1** | **A2** | **AF_A1** | **BETA** | **OR** | **SE** | **P** | **AF_A1** | **BETA** | **OR** |
| 6 | *HLA-DQB1* | rs9273364 | 32658525 | T | G | 0.64 | -1.53 | 0.22 | 0.08 | 3.03E-82 | 0.71 | -1.53 | 0.22 |
| 10 | *NRP1* | rs722988 | 33137219 | T | C | 0.50 | -0.48 | 0.62 | 0.08 | 1.10E-08 | 0.46 | -0.38 | 0.69 |
| 11 | *INS* | rs10770140 | 2172367 | T | C | 0.55 | 0.52 | 1.69 | 0.09 | 1.18E-09 | 0.51 | 0.49 | 1.64 |

| | | | | AMR | | | | | Pseudo case-control (mostly EUR) | | |
|---|---|---|---|---|---|---|---|---|---|---|---|
| **SE** | **P** | **AF_A1** | **BETA** | **OR** | **SE** | **P** | **AF_A1** | **BETA** | **OR** | **SE** | **P** |
| 0.31 | 0.1376 | 0.06 | 0.53 | 1.70 | 0.33 | 0.1071 | 0.13 | 0.51 | 1.66 | 0.05 | 1.14E-21 |
| 0.10 | 0.1238 | 0.52 | -0.48 | 0.62 | 0.16 | 0.0020 | 0.55 | -0.16 | 0.86 | 0.04 | 1.78E-05 |
| 0.09 | 1.42E-37 | 0.37 | -1.38 | 0.25 | 0.16 | 1.83E-18 | 0.39 | -1.30 | 0.27 | 0.03 | 8.44E-317 |
| 0.10 | 0.1351 | 0.59 | -0.25 | 0.78 | 0.18 | 0.1516 | 0.45 | -0.17 | 0.84 | 0.04 | 3.27E-06 |
| . | . | 0.04 | -1.13 | 0.32 | 0.41 | 0.0056 | 0.06 | -0.41 | 0.67 | 0.08 | 1.98E-07 |
| 0.10 | 0.0257 | 0.58 | -0.71 | 0.49 | 0.16 | 6.53E-06 | 0.58 | -0.13 | 0.88 | 0.04 | 0.0003 |
| 0.17 | 0.1071 | 0.14 | -0.62 | 0.54 | 0.26 | 0.0188 | 0.18 | -0.24 | 0.79 | 0.05 | 4.35E-07 |
| 0.11 | 0.0002 | 0.22 | -0.92 | 0.40 | 0.21 | 7.75E-06 | 0.22 | -0.62 | 0.54 | 0.05 | 2.41E-39 |
| 0.20 | 0.3731 | 0.22 | 0.33 | 1.40 | 0.20 | 0.0960 | 0.33 | 0.27 | 1.31 | 0.04 | 4.02E-12 |
| 0.18 | 0.3978 | 0.27 | 0.42 | 1.52 | 0.18 | 0.0175 | 0.48 | 0.23 | 1.26 | 0.04 | 6.32E-10 |
| 0.10 | 0.1093 | 0.55 | -0.04 | 0.96 | 0.17 | 0.8135 | 0.47 | -0.18 | 0.84 | 0.04 | 1.33E-06 |

| | | | | AMR | | |
|---|---|---|---|---|---|---|
| **SE** | **P** | **AF_A1** | **BETA** | **OR** | **SE** | **P** |
| 0.10 | 7.93E-58 | 0.48 | -1.53 | 0.22 | 0.14 | 3.42E-26 |
| 0.10 | 0.0001 | 0.60 | -0.75 | 0.47 | 0.16 | 2.92E-06 |
| 0.10 | 4.34E-07 | 0.69 | 0.63 | 1.88 | 0.18 | 0.0006 |

**Supplementary Table 3.** Seven HLA class II haplotypes are independently associated with T1D in African-ancestry individuals

| DRB1 | DQA1 | DQB1 | OR | P |
|---|---|---|---|---|
| 03:01 | 05:01 | 02:01 | 5.60 | 3.00E-20 |
| 04:05 | 03:01 | 03:02 | 13.03 | 2.50E-10 |
| 04:01 | 03:01 | 03:02 | 9.83 | 3.80E-12 |
| 09:01 | 03:01 | 02:01 | 5.04 | 1.10E-10 |
| 07:01 | 03:01 | 02:01 | 10.70 | 2.20E-07 |
| 15:03 | 01:02 | 06:02 | 0.26 | 1.10E-04 |
| 11:01 | 01:02 | 06:02 | 0† | 9.80E-01 |

Odds ratios (OR) and P-values generated by multivariable logistic regression of T1D risk, including 4 principal components and all 7 alleles as independent

**Supplementary Table 4.** Two HLA class II haplotypes are independently associated with T1D in Admixed-ancestry individuals

| DRB1 | DQA1 | DQB1 | OR | P |
|---|---|---|---|---|
| 03:01 | 05:01 | 02:01 | 6.01 | 4.50E-07 |
| 08:02 | 04:01 | 04:02 | 0.39 | 2.90E-02 |

Odds ratios (OR) and P-values generated by multivariable logistic regression of T1D risk, including 4 principal components and 2 alleles as independent

**Supplementary Table 5.** Nineteen HLA class II haplotypes are independently associated with T1D in European-ancestry individuals

| DRB1 | DQA1 | DQB1 | OR | P |
|---|---|---|---|---|
| 04:01 | 03:01 | 03:02 | 8.09 | 2.10E-117 |
| 03:01 | 05:01 | 02:01 | 2.97 | 1.30E-52 |
| 15:01 | 01:02 | 06:02 | 0.05 | 3.70E-40 |
| 04:04 | 03:01 | 03:02 | 3.15 | 3.00E-21 |
| 04:02 | 03:01 | 03:02 | 3.83 | 3.70E-17 |
| 04:05 | 03:01 | 03:02 | 6.36 | 6.50E-17 |
| 11:01 | 05:01 | 03:01 | 0.22 | 1.80E-18 |
| 13:01 | 01:03 | 06:03 | 0.25 | 5.60E-18 |
| 11:04 | 05:01 | 03:01 | 0.11 | 2.80E-13 |
| 07:01 | 02:01 | 03:03 | 0.13 | 7.30E-10 |
| 14:01 | 01:01 | 05:03 | 0.05 | 7.80E-07 |
| 04:05 | 03:01 | 02:01 | 6.4 | 1.10E-05 |
| 08:01 | 04:01 | 04:02 | 1.81 | 5.50E-05 |
| 04:07 | 03:01 | 03:01 | 0.09 | 8.10E-05 |
| 07:01 | 02:01 | 02:01 | 0.55 | 8.80E-09 |
| 15:02 | 01:03 | 06:01 | 0.16 | 2.80E-04 |
| 13:03 | 05:01 | 03:01 | 0.24 | 3.10E-04 |
| 10:01 | 01:01 | 05:01 | 0.29 | 9.60E-04 |
| 12:01 | 05:01 | 03:01 | 0.43 | 1.70E-03 |

Odds ratios (OR) and P-values generated by multivariable logistic regression of T1D risk, including 4 principal components and all 19 alleles as independent variables.
