## Supplemental Figures 1-4 for "A multi-ancestry genome-wide association study in type 1 diabetes"

### Supplementary Figures

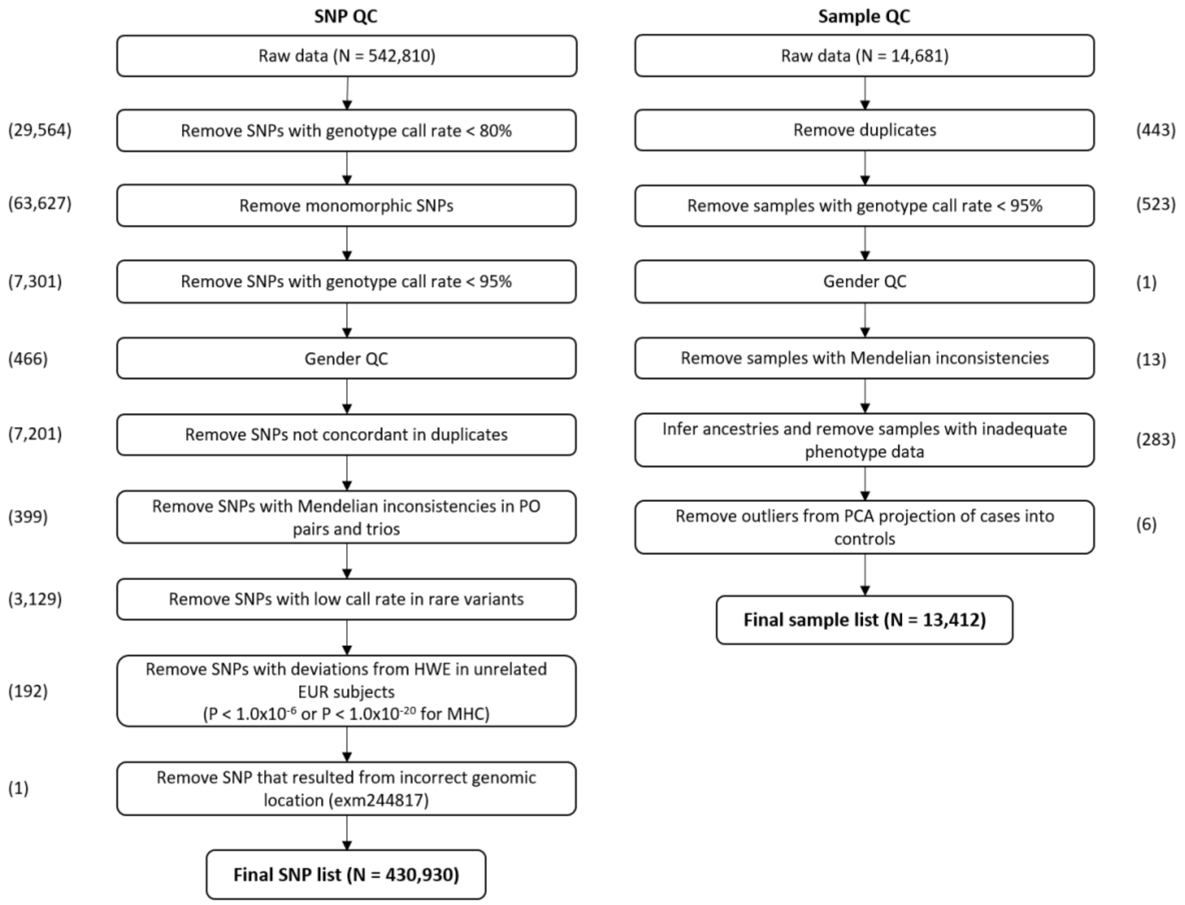


***Supplementary Figure 1.*** *SNP and sample quality control workflow before imputation.*

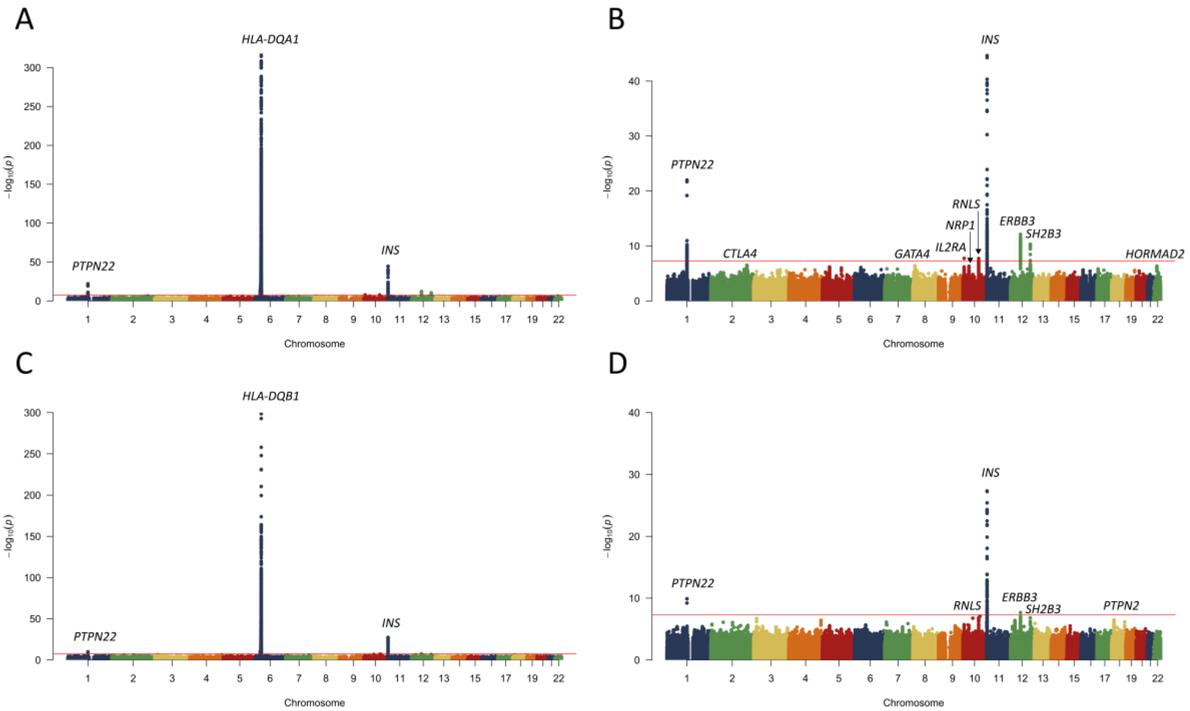


**Supplementary Figure 2.** Loci associated with type 1 diabetes risk (**A, B**) and age at onset (**C, D**) in AFR, AMR and pseudo case-control meta-analysis. Each locus is labeled with the nearest gene. The y-axis represents –log(P values). The horizontal red line represents the threshold for genome-wide associations. (**A**) The strongest associations with type 1 diabetes risk (*HLA-DQA1*, *INS* and *PTPN22*). (**B**) Associations with type 1 diabetes risk after excluding HLA region (25 Mb – 35 Mb). (**C**) The strongest associations with age at onset of type 1 diabetes (*HLA-DQA1*, *INS* and *PTPN22*). (**D**) Associations with age at onset of type 1 diabetes after excluding HLA region (25 Mb – 35 Mb).

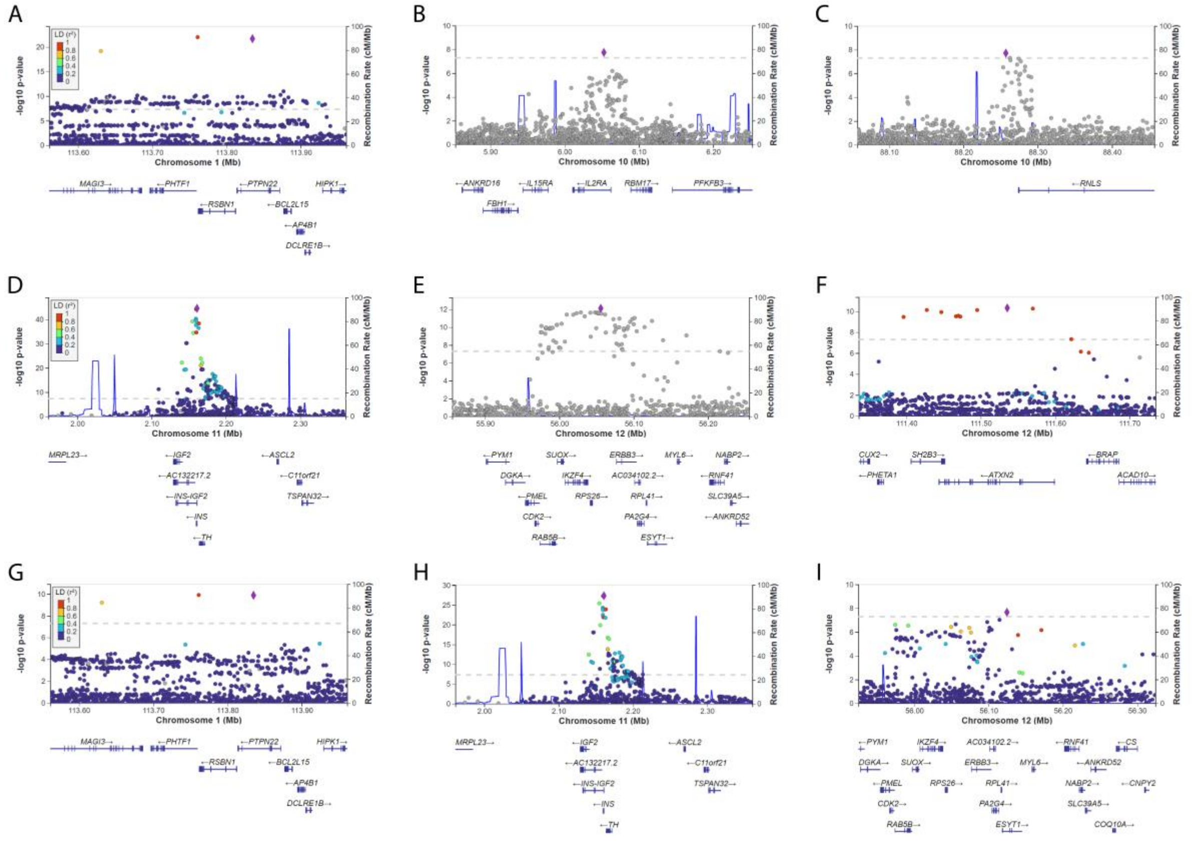


**Supplementary Figure 3.** LocusZoom plots of genome-wide significant loci associated with type 1 diabetes risk (**A-F**) and age at onset (**G-I**), excluding HLA region. The left y-axis represents –log(P values) and right y-axis determines recombination rate. The horizontal gray dashed line represents the threshold for genome-wide associations. The most significantly associated SNP is shown as purple diamond. For *PTPN22* locus (**A, G**), the purple diamond represents known coding region variant.

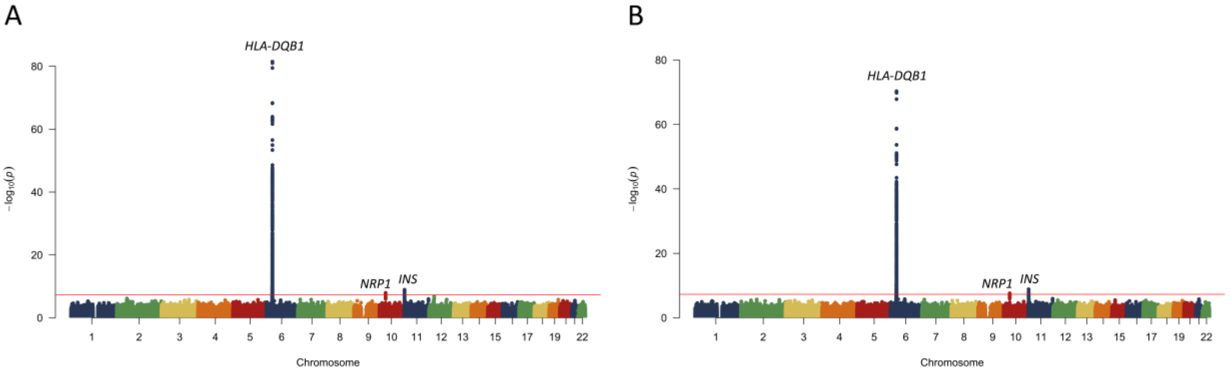


**Supplementary Figure 4.** Loci associated with type 1 diabetes risk (**A**) and age at onset (**B**) in non-EUR meta-analysis (AFR and AMR). Each locus is labeled with the nearest gene. The y-axis represents –log(P values). The horizontal red line represents the threshold for genome-wide associations. (**A**) Associations with type 1 diabetes risk in non-EUR meta-analysis. (**B**) Associations with age at onset of type 1 diabetes in non-EUR meta-analysis.
